# Effects of Tocilizumab in Critically Ill Patients With COVID-19: A Quasi-Experimental Study

**DOI:** 10.1101/2020.07.13.20149328

**Authors:** Victor Carvalho, Ricardo Turon, Bruno Gonçalves, Victor Fraga Ceotto, Pedro Kurtz, Cássia Righy

## Abstract

**Objectives:** Critically ill patients with COVID-19 may suffer from a cytokine release syndrome (CRS) characterized by remarkably high levels of interleukin 6 (IL-6). We assessed the effects of tocilizumab, an IL-6 receptor antagonist, on intra-hospital mortality and development of positive cultures in patients with COVID-19 admitted to the ICU.

**Design:** Patients with COVID 19 admitted in the ICU who were treated with tocilizumab plus standard care were enrolled and compared to controls.

**Setting:** COVID-19 severe disease

**Patients:** Patients with severe COVID-19 disease admitted in the ICU.

**Interventions:** Tocilizumab 400 mg IV two doses. Standard and intensive medical care as per institutional clinical protocol.

**Measures and Main Results:** Main outcome: 1) intra-hospital mortality; Secondary Outcomes: 1) the need for renal replacement therapy, 2) use of antibiotics and positive culture, and 3) inflammatory and oxygenation markers. There was no difference in mortality, need for renal replacement therapy, use of antibiotics or positive cultures between the two groups. The use of corticosteroids was more frequent in the treatment group. Levels of C-reactive protein (CRP) and WBC (white blood cells) counts declined significantly faster in the treatment group. Oxygenation markers rose significantly higher in patients in the tocilizumab group as compared to controls.

**Conclusion:** Tocilizumab was associated with rapid improvement in oxygenation and a faster decrease of CRP and WBC counts in patients with COVID-19 and should be evaluated as rescue therapy for patients with progressive disease

## Introduction

In 2019, an outbreak of a new virus known as SARS-CoV-2 has spread into a pandemic. On July 2020, Brazil has been the second most hit country in the world, accounting for almost 2 million cases. (1) The COVID-19 disease may vary from asymptomatic disease to a multiorgan systemic failure syndrome. Some studies have shown an association between patterns of cytokine release syndrome (CRS) with remarkably high levels of interleukin 6 (IL-6) and worse outcome in critically ill patients. (2,3) Tocilizumab is a recombinant humanized monoclonal antibody which antagonizes the IL-6 receptor. (4) Preliminary reports suggested that the use of tocilizumab in patients SARS-CoV-2 was associated with significant clinical and laboratory improvement. (5,6) This drug is not already labeled for use in this disease.

We evaluated the effect of tocilizumab on the intra-hospital mortality and other clinical and biomarker parameters of critically ill patients with COVID-19, as compared to controls. We also examined temporal changes in the inflammatory response of treated patients.

## Material and Methods

Patients with COVID-19 disease who met the inclusion criteria and were admitted to the ICU of our institution between 21st March and 31st May 2020 were recruited. All the patients received standard intensive medical care, as per institutional clinical protocol which includes hydroxychloroquine and azithromycin. Physicians were also able to use corticosteroids according to specific clinical indications (eg. acute respiratory distress syndrome). In addition, arterial blood gases analysis, complete blood count, CRP, lactic dehydrogenase, d-dimer, ferritin, and inflammatory biomarkers were collected daily.

### Study Design and Patients

This is a single center study. All the patients admitted to the ICU with suspected COVID-19 underwent diagnostic testing for SARS-CoV-2 through nasopharyngeal swabs. The patients admitted to the ICU also received respiratory support with either oxygen through a nonrebreather mask or non-invasive or mechanical ventilation. Patients with acute respiratory distress syndrome (ARDS) were managed with neuromuscular blockade and a protective ventilation strategy that included low tidal volume (6 mL/kg of predicted body weight) and driving pressure (less than 15 cmH2O) as well as optimal PEEP calculated based on the best lung compliance. In those with moderate to severe ARDS and P/F ratio below 150, despite optimal ventilatory settings, prone position was initiated.

After IRB approval and availability of tocilizumab, all consecutive adult patients (age > 18 years) admitted to the ICU with suspected or confirmed SARS-CoV-2 infection plus fever (axillary temperature > 38°C) or elevated CRP (≥ 5 mg/dL) and ventilatory or oxygenation deterioration or need for ventilatory support (non-invasive or mechanical ventilation) received two doses of intravenous tocilizumab 400 mg. Patients treated with tocilizumab were compared to controls admitted to our ICU before tocilizumab availability. Follow-up continued through June 7th or a minimum of 14 days for all patients.

The study protocol was approved by the National Ethics Committee (4.059.207), who waived the need for informed consent. The study protocol was registered at the Brazilian Registry of Clinical Trials (ReBEC) with the identification RBR-3zdynp.

### Exclusion criteria

Patients with primary or secondary immunodeficiency, using immunosuppressive drugs, pregnant women, those without respiratory symptoms and in palliative care.

### Main Outcomes

Main outcomes analyzed were intra-hospital mortality, need for renal replacement therapy, positive cultures and use of antibiotics

### Statistical Analysis

Univariate associations were tested by using the chi-square or Fisher’s exact test for categorical variables, the two-tailed t-test for normally distributed continuous variables, and the Mann–Whitney U test for non-normally distributed continuous variables. Multivariable analysis was performed to determine if there was any association of the variables of the study and mortality or positive microbiological sample.

Non-linear logistic regression was used to test the difference between trends and slopes of the curve of the oxygenation markers (P/F ratio and SpO2/FiO2 ratio) and of inflammatory biomarkers (C-reactive protein and white blood cells count). The daily progression was assessed from the day of the start of tocilizumab in the treatment group (index day) through day five, and from the 3rd day after admission on the control group through day seven - the 3rd day was chosen since it is the median day of the start of tocilizumab in the treatment group. To make the analysis of the slope more precise, P/F ratio and SpO2/FiO2 ratio were normalized by dividing each daily value by the index day, which made possible to analyze the actual trend. The significance was set to 0.05 for all analyses. Statistical analyses were performed using IBM SPSS Statistics version 23.0 (Chicago, IL, USA) and Prism (GraphPad, San Diego, CA, USA)

## Results

### Baseline characteristics

Fifty-three patients were enrolled in the study, twenty-nine in the treatment group and twenty-four in the control group. The median age was 55 (44-65) years in the tocilizumab group and 58.5 (51-70.8) years in the control group and most patients were male (62% × 75%, respectively). Corticosteroid use was more frequent in the treatment group (83% × 37%, p=0.001). The clinical and demographic characteristics on admission are detailed in Table 1. In-hospital mortality was 17% in both groups.

**Table 1.** Clinical and demographic characteristics on admission (values in median + interquartile range and n + percentage)

| variable | tocilizumab (n = 29) | control ( n = 24) | p-value |
| --- | --- | --- | --- |
| age, y | 55 (44-65) | 58.5 (51-70.8) | 0.34 |
| female gender | 11 (38%) | 6 (25%) | 0.38 |
| saps III on admission | 43 (39-55) | 45 (38.8-54) | 0.87 |
| days of symptom onset to admission | 6 (5-8) | 8 (5-10) | 0.18 |
| mechanical ventilation | 15 (52%) | 7 (29%) | 0.16 |
| corticosteroids use | 24 (83%) | 9 (37%) | 0.001 |
| hct + azt use | 22 (76%) | 17 (71%) | 0.76 |
| **wbc/mm<sup>3</sup>** | 7200<br>(5700-9500) | 6500<br>(5375-9575) | 0.77 |
| **cpr (mg/dL)** | 19.5<br>(8.18-28.2) | 15.8<br>(6.28-20.3) | 0.08 |
| **ldh U/L** | 497<br>(379.8-656.5) | 374<br>(291-511) | 0.05 |
| **d-dimer ng/mL** | 1128.5<br>(518-1501.7) | 1514.5<br>(798.5-2470.5) | 0.09 |
| **ferritin ng/mL** | 1124.5<br>(644.3-1539.7) | 871.7<br>(432.8-1509.6) | 0.49 |

*crp* – c-reactive protein; *hcq + azt* – hydroxychloroquine plus azithromycin; *ldh* – lactate dehydrogenase; *saps* – simplified acute physiology score; *wbc* – white blood cells.

### Outcomes: univariate and multivariable analysis

In the univariate analysis, tocilizumab was not associated with any of the outcomes assessed: mortality, positive cultures, use of antibiotics or need for renal replacement therapy (Table 2). In the multivariable analysis, after adjusting for age and mechanical ventilation, use of tocilizumab was not associated with mortality (OR 3.97, 95% CI 0.28-57.2, p=0.3) or positive cultures (OR 1.73, 95% CI 0.22-13.82, p=0.6) (Table 3). No adverse events were reported that could be directly related to the administration of tocilizumab.

**Table 2.** Primary and secondary outcomes – Univariate analysis (values in n + percentage)

|  | **Tocilizumab** | **Control** | **Univariate analysis** |
| --- | --- | --- | --- |
|  | n = 29 | n = 24 | p-value |
| **mortality** | 5 (17%) | 4 (17%) | 1 |
| **any positive culture** | 11 (38%) | 4 (17%) | 0.13 |
| **bacterial positive culture** | 10 (34%) | 3 (12%) | 0.1 |
| **fungal positive culture** | 6 (21%) | 1 (4%) | 0.12 |
| **use of ATB** | 19 (66%) | 15 (63%) | 1 |
| **need of RRT** | 9 (31%) | 3 (13%) | 0.18 |

*ATB – antibiotics; RRT – renal replacement therapy.*

**Table 3.** Multivariable analysis – Outcome: positive culture

|  | **OR** | **95% CI** | **p-value** |
| --- | --- | --- | --- |
| **age over 60** | 0.13 | 0.01-1.57 | 0.1 |
| **female gender** | 10.22 | 0.78-134.62 | 0.08 |
| **mechanical ventilation** | 256.47 | 5.1-12899.74 | 0.006 |
| **use of tocilizumab** | 1.73 | 0.22-13.82 | 0.6 |
| **use of corticosteroids** | 0.3 | 0.02-4.93 | 0.4 |

### Daily progression of inflammatory and gas exchange markers

Though not different at baseline, on the index day, the tocilizumab group had both higher inflammatory response markers (median CRP 20.8 mg/dL x 13.5 mg/dL in the control group, p=0.0005) and lower gas exchange ratio (165 x 264 in the control group, p=0.000, as seen in Table 4 (daily progression of variables)

The CRP curve slope (Figure 2) is significantly different in the two groups, with a markedly decrease in the tocilizumab group (p=0.009), as well as the WBC curve slope (p=0.02) (Figure 3). The slope of the rise of oxygenation, in normalized values, is also significantly different, being higher in the tocilizumab group (p=0.02) (Figure 1).

**Table 4.** Daily progression of variables (values in median + interquartile range)

|  |  | **D1 (index)** | **D2** | **D3** | **D4** | **D5** |
| --- | --- | --- | --- | --- | --- | --- |
| **crp mg/dL** | **tocilizumab** | 20.8 (15.5-36) | 22.4 (15.18-28.15) | 14.3 (5.5-22.3) | 4.75 (3.08-8.1) | 2.8 (2.1-5.2) |
|  | **control** | 13.5 (5-17.25) | 7 (5.25-8.4) | 4.9 (3.8-11.3) | 3.5 (2.75-15.63) | 5 (2.13-7.75) |
|  | **p-value** | 0.0005 | 0.001 | 0.023 | 0.44 | 0.07 |
| **wbc/mm3** | **tocilizumab** | 9600 (7350-14650) | 8500 (5600-12700) | 7300 (4800-11000) | 8700 (6800-11850) | 10100 (6400-14650) |
|  | **control** | 6900 (5000-9025) | 7500 (5350-10200) | 6100 (5300-9150) | 7050 (5375-13125) | 7900 (6625-15525) |
|  | **p-value** | 0.08 | 0.87 | 0.45 | 0.8 | 0.9 |
| **pao2/fio2 and**<br>**sao2/fio2**<br>**ratio** | **tocilizumab** | 165<br>(124-251) | 204<br>(144-288) | 274<br>(185-371) | 227<br>(188-308) | 283<br>(213-380) |
|  | **control** | 264<br>(203-420) | 300<br>(183-420) | 346<br>(227-420) | 420<br>(206-420) | 342<br>(267-420) |
|  | **p-value** | 0.0002 | 0.075 | 0.36 | 0.04 | 0.15 |
| **normalized**<br>**pao2/fio2 and**<br>**sao2/fio2**<br>**ratio** | **tocilizumab** | 1 | 1.11 (0.91-1.41) | 1.42 (0.89-1.82) | 1.2 (1.04-1.53) | 1.25 (1.17-1.72) |
|  | **control** | 1 | 1 (0.98-1.04) | 1 (0.91-1.23) | 1 (0.95-1.59) | 1.06 (0.99-1.53) |
|  | **p-value** | - | 0.043 | 0.02 | 0.14 | 0.07 |

*crp* – c-reactive protein; *wbc* – white blood cells.

**Figure 1.**
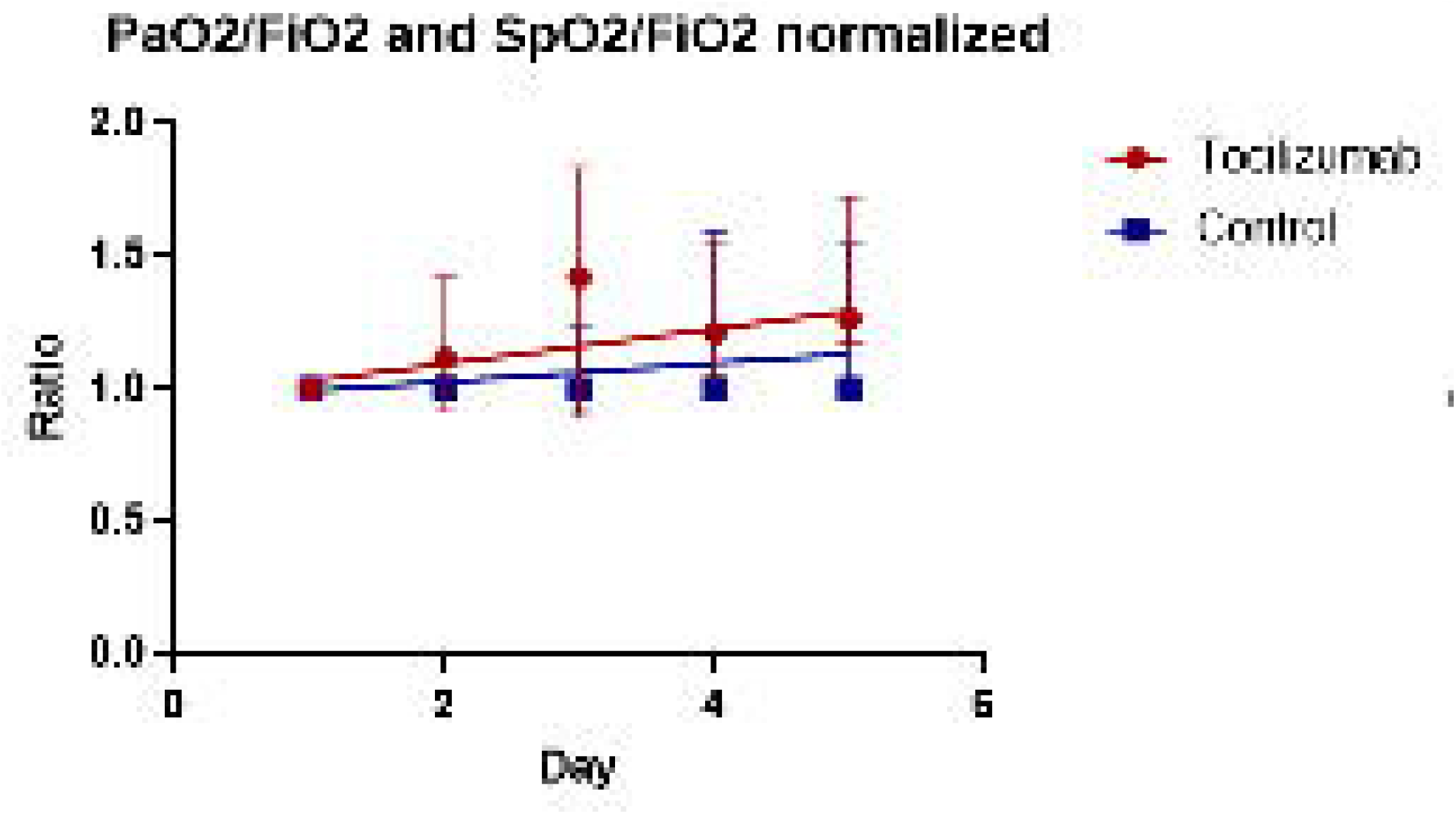
Nonlinear regression of daily progression of normalized PaO2/FiO2 and SpO2/FiO2 ratio (values in median + interquartile range); p=0.02.

**Figure 2.**
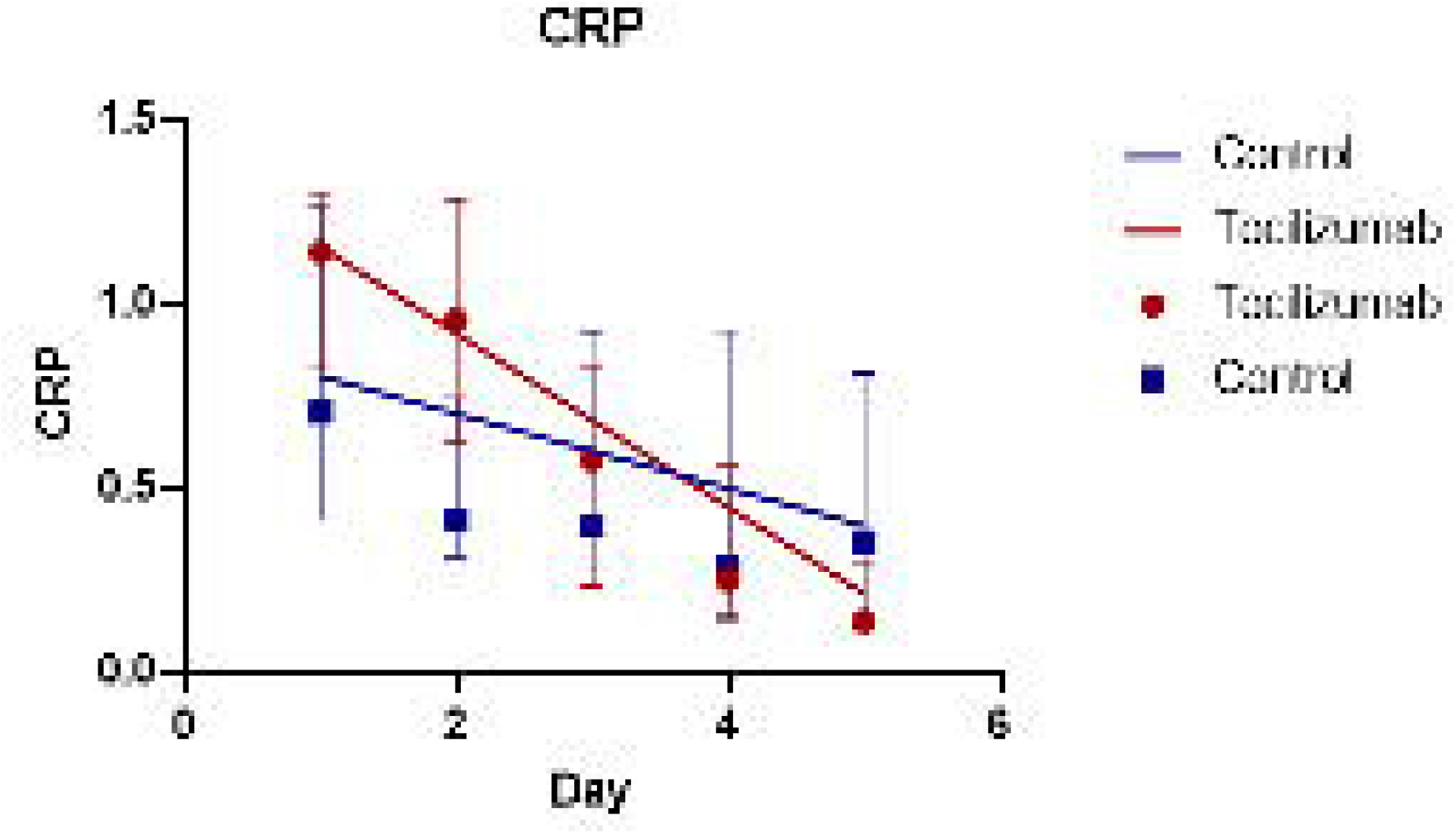
Nonlinear regression of daily progression of normalized C-reactive protein (CRP) in mg/dL (values in median + interquartile range); p=0.009.

**Figure 3.**
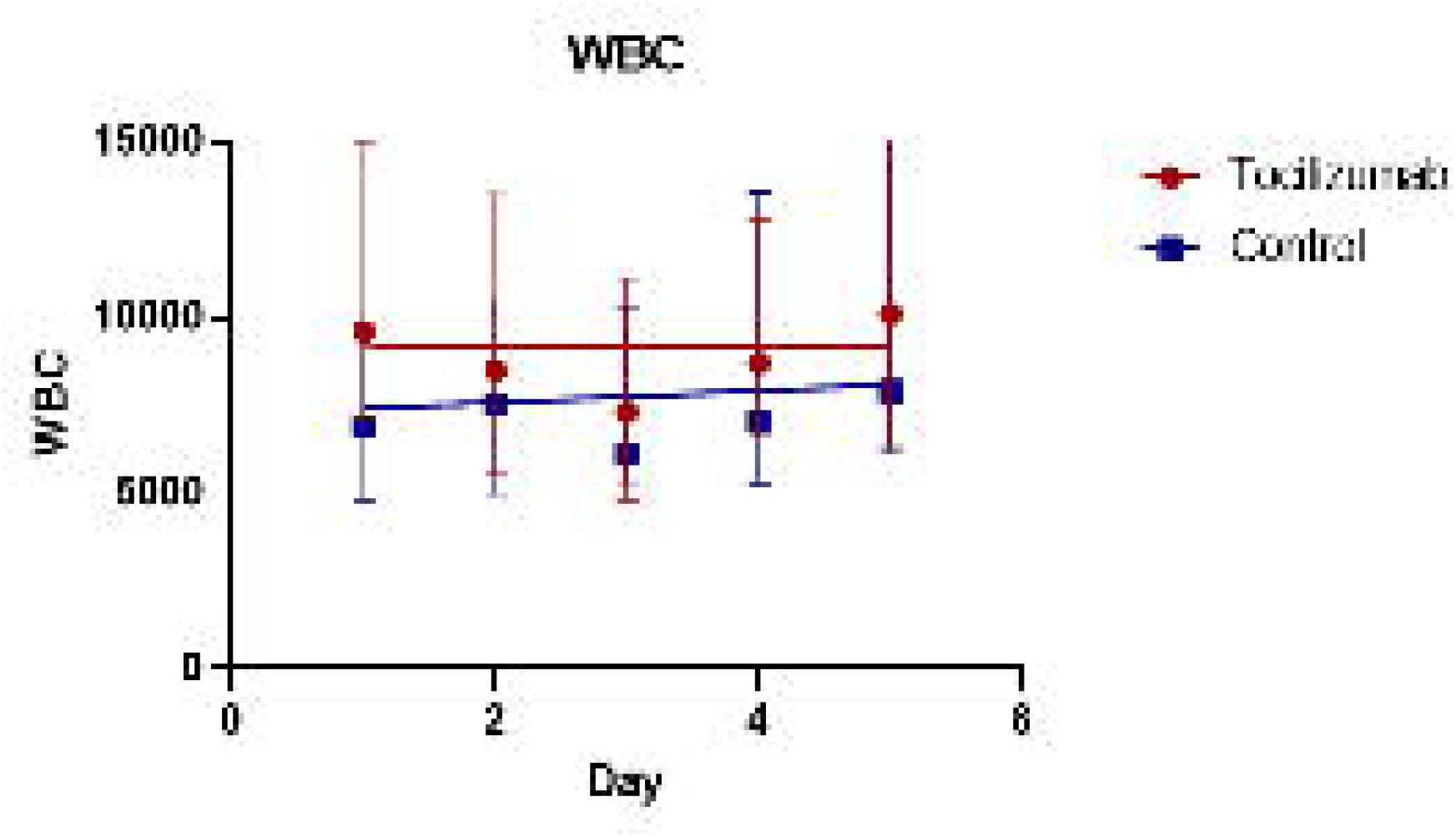
Nonlinear regression of daily progression of white blood cells count (WBC)/mm3 (values in median + interquartile range); p=0.02.

## Discussion

Although patients in the tocilizumab group were more severely ill, there was no difference neither in-hospital mortality nor secondary infections. Furthermore, similarly to other studies (5-9), use of tocilizumab was associated with a faster improvement of oxygenation (slope of normalized P/F and SpO2/FiO2 Ratio; p = 0.02) and a decrease of CRP levels (slope of the CRP curve; p = 0.009).

Considering that Tocilizumab and corticosteroid tend to reduce the inflammation and immune response, and these effects could last for weeks, it is reasonable to expect a higher number of infections in the tocilizumab group.(6) There was a non-significant increase in the number of positive cultures in the treatment group (38% vs 17%), although there was no difference either in the use of antibiotics or in clinical changes suggesting secondary infections such as the increased need for vasoactive amine, worsening of the PaO2/FiO2 ratio, changes in pulmonary secretion (quantity and quality), or signs of inflammation on catheter sites. Moreover, just one patient (3.4%) developed refractory septic shock and died. These results are similar to an Italian cohort, which reported two deaths by septic shock (2%) during the 10-day follow-up (7). In this sense, the number of positive cultures in the intervention group suggests that these patients possibly are more susceptible only to colonization and not secondary infections, as expected.

Therefore, the improvement in oxygenation and inflammatory markers with no increase of infections suggests that tocilizumab could be an option as rescue therapy for patients with progressive COVID-19 after initiation of systemic dexamethasone, as shown in RECOVERY trial (10). Furthermore, the administration of tocilizumab seems to reduce ICU admission, making, along with dexamethasone, an option of treatment in selected cases of critically ill patients (8). However, our study has some limitations. Corticosteroid use was not balanced between groups, limiting the determination of the real benefit in oxygenation and CRP reduction of tocilizumab in patients who used corticosteroid; the number of patients was limited; it was a single center study, neither blind nor randomized. Nevertheless, this study generates an interesting hypothesis on the safety of concomitant use of tocilizumab and corticosteroid in COVID patients. This hypothesis should be evaluated in a larger clinical trial.

## Conclusions

The improvement in oxygenation and inflammatory markers with no increase secondary infections or intra-hospital mortality suggests that tocilizumab could be an option for patients with progressive COVID-19 after initiation of systemic corticosteroids.

## Data Availability

I will make the data and associated documentation available to users under a data-sharing agreement.

